# Giving birth in a Pandemic: Women’s Birth Experiences in England during COVID-19

**DOI:** 10.1101/2021.07.05.21260022

**Authors:** Ezra Aydin, Kevin A. Glasgow, Staci M. Weiss, Zahra Khan, Topun Austin, Mark H. Johnson, Jane Barlow, Sarah Lloyd-Fox

## Abstract

**Background:** Expectant parents worldwide have experienced changes in the way they give birth as a result of COVID-19, including restrictions relating to access to birthing units and the presence of birthing partners during the birth, and changes to birth plans. This paper reports the experiences of women in England.

**Methods:** Data were obtained from both closed- and open-ended responses collected as part of the national COVID in Context of Pregnancy, Infancy and Parenting (CoCoPIP) Study online survey (n = 477 families) between 15^th^ July 2020 – 29^th^ March 2021. Frequency data are presented alongside the results of a sentiment analysis; the open-ended data was analysed thematically.

**Results:** Two-thirds of expectant women reported giving birth via spontaneous vaginal delivery (SVD) (66.1%) and a third via caesarean section (CS) (32.6%) or ‘other’ (1.3%). Just under half (49.7%) of the CS were reported to have been elective/planned, with 47.7% being emergencies. A third (37.4%) of participants reported having no changes to their delivery, with a further 25% reporting COVID-related changes, and 37.4% reporting non-COVID related changes (e.g., medical intervention). Experiences of COVID-related changes included limited birthing options and reduced feelings of control; difficulties accessing pain-relief and assistance, and feelings of distress and anxiety. Under half of the respondents reported not knowing whether there could be someone present at the birth (44.8%), with 2.3% of respondents reporting no birthing partner being present due to COVID-related restrictions. Parental experiences of communication and advice provided by the hospital prior to delivery were mixed, with significant stress and anxiety being reported in relation to both the fluctuating guidance and lack of certainty regarding the presence of birthing partners at the birth. The sentiment analysis revealed that participant experiences of giving birth during the pandemic were predominately negative (46.9%) particularly in relation to the first national lockdown, with a smaller proportion of positive (33.2%) and neutral responses (19.9%).

**Conclusion:** Parents reported an overall increase in birthing interventions (e.g., emergency CS), increased uncertainties related to the birth, and poor communication, leading to increased feelings of anxiety and high levels of negative emotions. The implications of these findings are discussed.

## Introduction

In January 2020 the first case of COVID-19 in the UK was confirmed, and on the 23^rd^ of March 2020 a national lockdown was announced. For this and two later national lockdowns in England, all non-essential businesses were closed, and people were required to stay at home, being permitted to leave for essential purposes only (e.g., medical workers) (see *Figure 1* for timeline and dates of restrictions and guidelines in England).

**Figure 1:**
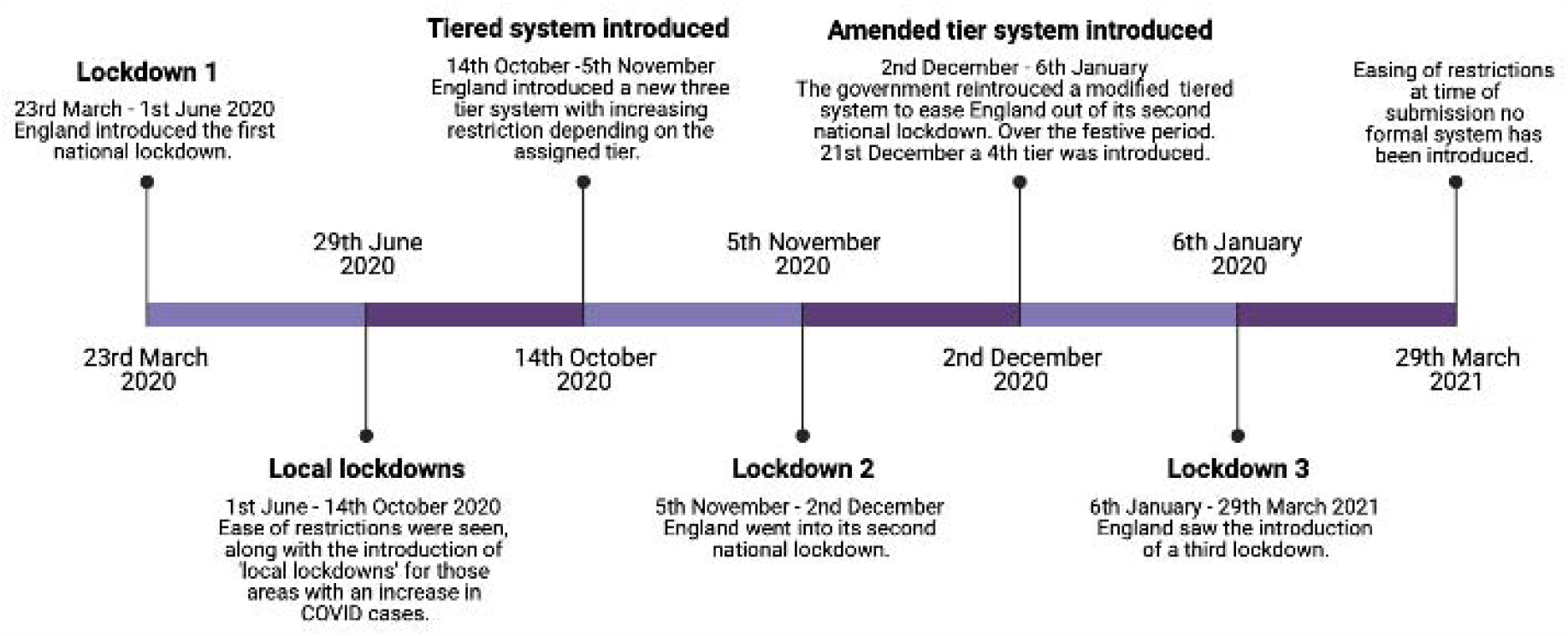
Timeline of restrictions and guidelines* imposed by the government between March 2020-March 2021 *It is important to note that some areas in England may have seen a slight alteration in between national guidance and restrictions rules in his time period.

Throughout the pandemic, pregnancy and childbirth have been associated with anxiety and uncertainty for many pregnant women and their partners due in part to the changing landscape of the healthcare system and increased demands on healthcare providers. This has resulted in a number of best practices endorsed by the World Health Organising (WHO) being side-lined as evidenced by reports of women giving birth alone (San Francisco: Human Rights in Childbirth, 2020; Walsh et al., 2020), restrictions being imposed on birthing options (e.g., no water births) (Greenfield et al., 2021; Nelson & Romanis, 2020)), and separation from their baby shortly after birth (San Francisco: Human Rights in Childbirth, 2020).

In the UK, government guidelines aimed at curbing the spread of the virus also led to a number of suboptimal conditions for expectant parents giving birth. In March 2020, National Health Service (NHS) trusts began to suspend home birth services and support in response to the COVID-19 outbreak (Davis, 2020). This was the result of a shortage in the number of midwives and maternity support workers (i.e., The Royal College of Midwives (RCM) reported a doubling in the shortage of midwives since the start of the COVID-19 outbreak - Royal College of Midwives, 2020; Sherwood, 2020), the diversion of resources to the pandemic, and ambulance shortages. Due to the suspension of NHS-supported home birth services, the RCM reported a surge in expectant women removing themselves from NHS antenatal care and a spike in private midwifery services, with increased numbers of expectant parents avoiding routine and obstetric care in hospitals (Davis, 2020). One study showed that between April and July 2020, one in 20 expectant women were considering giving birth without a doctor or midwife present (‘freebirth’) in the UK, 3% higher than recorded in 2019 (Greenfield et al., 2021). This qualitative study attributed the increased demand for ‘freebirths’ to wanting to avoid hospitals, fewer choices in terms of birth preferences (e.g., having a birthing partner present), and practical problems (e.g., inability to use public transport) (Greenfield et al., 2021; Nelson & Romanis, 2020).

From the beginning of the pandemic, individual NHS trusts were required to draw up their own guidance with regard to access to maternity services and birth partners, based on government guidelines. Most commonly, the guidance stated that partners were only allowed to be present when the mother was 4cm dilated, that they were not allowed to be present at the start of an induction and were not allowed to join their partners during the pre-operation preparation for a caesarean section (CS); and that they were to leave shortly after the birth (Regan, 2020). A survey of 15,000 new and expectant women conducted by the UK-based charity ‘Pregnant then Screwed’ between 16-18^th^ July 2020 found that 90% reported hospital restrictions to have had a negative impact on their mental health, with 97% reporting these restrictions to have also increased their anxieties related to childbirth. Furthermore, just under a fifth (17.4%) of respondents reported feeling ‘forced’ to have a vaginal examination whilst in labour with 82% feeling this was a requirement if they were to have their birthing partner join them during the delivery.

On the 8^th^ September 2020, NHS England issued guidance to individual NHS trusts “to reintroduce access for partners, visitors and other supporters of pregnant women in English maternity services” (Royal College of Obstetricians and Gynaecologists et al., 2020). However, the Guardian reported that only around 23% of trusts during this period allowed partners to be in attendance for the duration of the labour (Topping & Duncan, 2020), suggesting this guidance was applied inconsistently across trusts. In December 2020 this guidance was further revised to explicitly allow in-person support for expectant women throughout their maternity journey. This was inclusive of antenatal visits, ultrasound scans, and during the birth (NHS, 2020).

The COVID-19 in the Context of Pregnancy, Infant Parenting (CoCoPIP) Study was developed to explore how COVID-19 and the cascade of changes in healthcare, social restrictions and government guidance impacted the lives of families who were expecting a baby or had recently given birth (Aydin, Weiss, et al., 2021). Previously, data collected from the Context of Pregnancy, Infancy and Parenting (CoCoPIP) study was used to qualitatively explore expectant family’s perceptions of their healthcare appointments, health and social support in the UK during the pandemic (Aydin, Glasgow, et al., 2021). The aim of the current analysis was to explore parent’s experiences of giving birth during COVID-19, including the ways in which communication and advice provided by hospitals may have influenced these experiences.

## Methods

### Participants

Survey data was taken from the period 15^th^ July 2020 – 29^th^ March 2021 (n = 477, see *Table 1* for demographic and birth information). Recruitment strategies included the distribution of information nationwide to antenatal and postnatal health groups, social media platforms (Twitter, Facebook and Instagram), as well as other child development research groups and networks in the UK. Eligibility criteria for the study included expectant parents past their first trimester, or parents of an infant between the ages of 0-6 months, who were then asked to report on experiences during their recent pregnancy. Women who gave birth prior to the first national lockdown (23^rd^ March 2020), were excluded from the final analysis. Additionally, due to the differences in timings with regard to the guidance issued across England, Wales and Scotland, only those families who lived in England at time of birth were included in the final sample. These were identified by the postcode participants provided at time of completing the survey. All participating parents gave informed consent to take part in the CoCoPIP online survey (tinyurl.com/CoCoPIP) (Aydin, Weiss, et al., 2021). Ethics approval for the survey was given by the University of Cambridge, Psychology Research Ethics Committee (PREC) (PRE.2020.077).

**Table 1:** Participant demographic information

| Demographic | n |
| --- | --- |
| **Who** |  |
| Mother | 436 |
| Father | 7 |
| Non-birth Mother | 1 |
| Other partner | 0 |
| Missing | 33 |
| **Ethnicity** |  |
| White | 408 |
| Black | 13 |
| Asian | 7 |
| Mixed/Multiple | 15 |
| Hispanic | 0 |
| Arab | 0 |
| Other | 1 |
| Undisclosed | 1 |
| **Gestational age at birth** |  |
| Very preterm (<32 weeks) | 3 |
| Moderately preterm (32 – 33 weeks + 6 days) | 8 |
| Late preterm (34 – 36 weeks + 6 days) | 21 |
| Early term (37 – 38 weeks + 6 days) | 100 |
| Full term (39 – 41 weeks and 6 days) | 321 |
| Post term (>42 weeks) | 24 |
| **Delivery method** |  |
| Spontaneous vaginal delivery (SVD) | 316 |
| C-section | 155 |
| *Elective* | 77 |
| *Emergency* | 74 |
| Other | 6 |

### Procedure

The CoCoPIP survey comprises a mixed-methods approach, in which both quantitative and qualitative data was collected. This survey is logic-dependent and adaptive, only showing questions relevant to the parent’s current situation (i.e., first trimester/second trimester/infant aged 0-3/3-6 months). For the full survey, response time was ∼30 minutes and respondents were included in a £100 gift card prize draw. As part of this survey, parents or caregivers were asked to complete structured questions about delivery type and whether their partner and/or family were present during the birth (see *Table 2* for questions), alongside semi-structured questions focussed on their experience of giving birth during a pandemic (see *Table 3* for questions).

**Table 2:** Structured questions about partner and/or family access to hospital during birth.

| Question |
| --- |
| Prior to the birth, were you certain whether partners and/or family could be present for the birth? |
| Yes |
| I wasn't sure |
| No |
| Missing |
| Was the partner and/or family present for the birth? |
| Yes |
| No |
| No due to COVID-19 restrictions |

**Table 3:** Semi-structured questions asked to participants regarding their birthing experiences

| Question |
| --- |
| Was the way you delivered your baby as you wanted to in your birth plan, or did it change? |
| Prior to the birth, were you certain whether partners and/or family could be present for the birth? If you like, let us know how this communication or advice from the hospital made you feel. |

### Analysis

Descriptive data is presented below in the form of frequencies. Quantitative analysis of the data involved a sentiment analysis. As with previous research (Aydin, Glasgow, et al., 2021), this was conducted manually. Responses to each question (see *Table 3*) were read and categorised as ‘positive’, ‘negative’ or ‘neutral’ by a single researcher (EA) and a cross check of 10% of the sentiment labels were conducted by a second researcher (KAG).

The qualitative data was imported from Qualtrics® via Redcap® (Harris et al., 2009) into NVivo 12 (QSR International) software. We adopted the same methodology used in previous qualitative research from the CoCoPIP study cohort (Aydin, Glasgow, et al., 2021). The coding trail was double checked by EA. Finally, confirmability was addressed by ensuring a clear presentation of participant responses, and by providing a clear rational for each step involved in the methods and analysis; furthermore, an additional researcher (KAG) conducted a reliability analysis of 25% of the data to confirm the themes and sub-themes identified. It is important to note that any data reported as occurring ‘as a direct result of COVID-19 only’ includes answers where the individual explicitly referenced ‘COVID’, ‘pandemic’ or ‘PPE use’ in their response to the semi-structured questions asked (see *Table 3*).

## Results

Of the 477 participants who responded to questions regarding their birthing experiences during the pandemic, a third completed the survey during one of the three national lockdowns (39.5%, 188), around half completed the survey during the period of easing of restrictions (51.5%, 245) and a small percentage were completed during the introduction of a tiered system (9%, 43) (see *Figure 1* for timeline).

In our sample two-thirds of expectant women gave birth via spontaneous vaginal delivery (SVD) (66.1%, 315), a third delivered via CS (32.6%, 155), and ‘other’ (1.3%, 6) (see *Table 1*). Of the 155 participants who reported giving birth by CS, just under half (49.7%, 77) reported having an elective/planned CS, with the remainder (47.7%; 74) having an emergency CS (4 participants did not report on whether their CS was an elective or emergency procedure).

A large proportion of respondents reported being uncertain about the restrictions relating to birthing partners (40.2%, 191), and a fifth reported that they were unaware prior to the birth whether birthing partners would be allowed to be present (14.9%, 71) (see *Table 2* for questions). At time of birth the majority of participants reported having their partner or a family member present (96.2%, 459), whilst a small number reported not being able to have anyone present at the birth (3.8%, 18) with 2.3% of these being due to COVID-19 related restrictions between 23^rd^ March 2020 - 29^th^ March 2021.

The results of the sentiment analysis showed that of the total responses across all questions (n = 706), 33.2% expressed positive, 19.9% neutral and 46.9% negative sentiments. When observing sentiment in relation to the governmental guidance and restrictions (*see Figure 1 for timeline*) participants responses consistently showed a higher negative sentiment towards their birthing experiences during the first national lockdown (56.9%), ease of governmental guidance and restrictions, (43%) and tiered guidance system (42.6%). Relative to these periods, during the second and third national lockdowns, participants showed an almost equal number of negative (34.2%) and neutral (36.8%) sentiments during lockdown 2 and a more positive sentiment (50%) with regard to their birthing experiences during lockdown 3 (see *Figure 2*).

**Figure 2:**
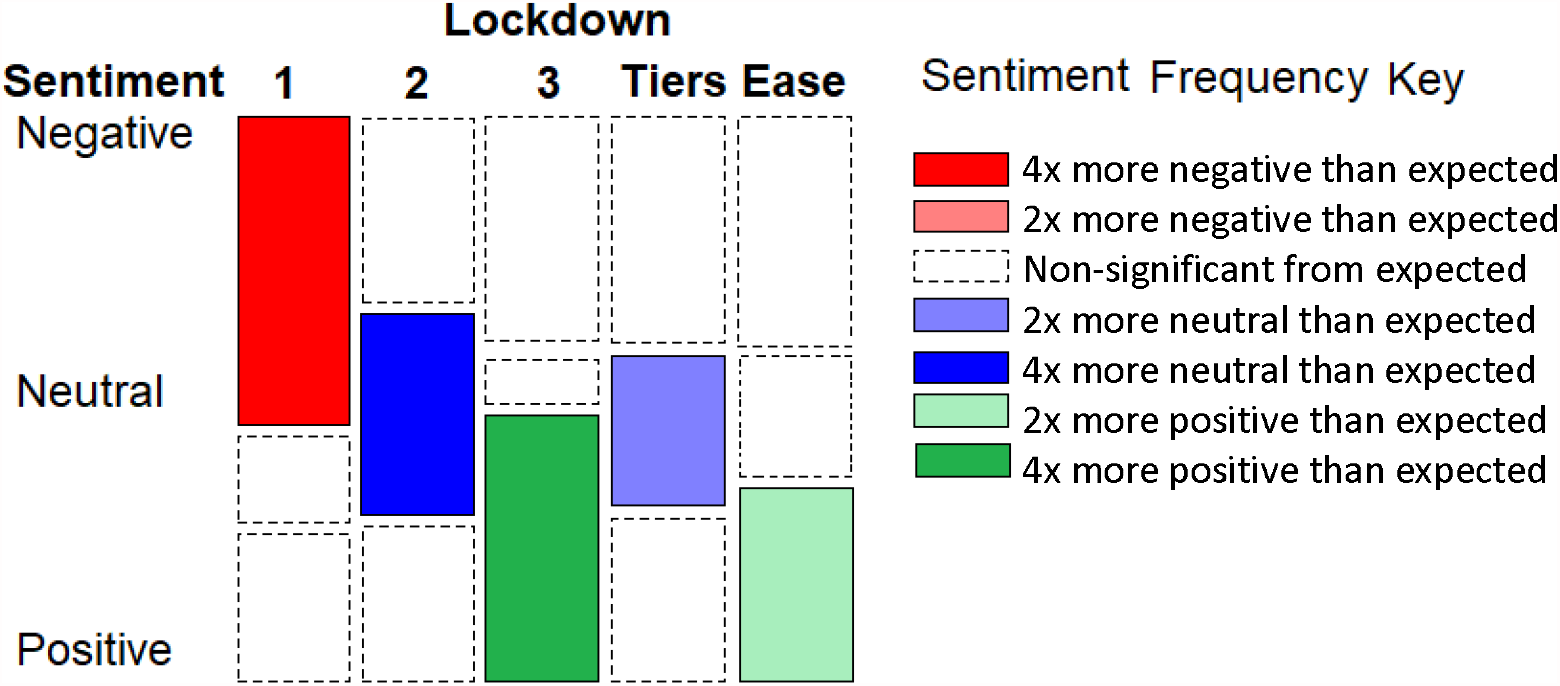
Diagram of sentiment analysis reported by national guidance group at time of birth.

Of the 462 respondents who responded to the question ‘*Was the way you delivered your baby as you wanted to in your birth plan, or did it change?* 37.4% (172) reported no changes to their planned delivery (although it should be noted that some of these responses suggested that no birth plan had been made as a result of the pandemic), 25% (115) reported changes to the planned delivery due to COVID and 37.4% (172) reported changes due to other reasons (e.g., changes in birth plan due to fetus being breech). Parents experiences of the COVID-related changes are described below:

### No changes to delivery plan

Although many parents across the UK experienced difficulties and hardships whilst giving birth, some respondents to the survey reported their birthing experience going according to their birthing plan, with parents recalling positive experiences in relation to the birth of their child:

> *‘My birthing experience was exactly how I’d planned/hoped. I had a very basic idea of how I’d like to give birth but was very open to other options. I was lucky enough to have a straight forward water birth with no complication’*

And

> *‘It was the way I wanted to. The birth experience was the most normal thing in the whole pregnancy’*

In addition to parents experiencing no changes to their birth plans, some participants described feeling supported and informed at the time of giving birth:

> *‘…. My birth plan was followed in that I was able to do and use the things I wanted and the staff knew that I was flexible should I need to be dependent on the situation that arose at the time*.*’*

Another participant referred to additional support offered due to previous birthing trauma:

> *‘Went exactly has[sic] planned and everyone was very calm and friendly due to previous birth trauma the year before*.*’*

However, a number of participants stated that there were no changes because birthing plans were not being made during national restrictions:

> *‘I was informed by my midwife that they were not making birthing plans during lockdown so I didn’t have a plan*.*’*

While this was experienced well by some women - *‘I had no birth plan so it all went how I would of liked it’-*other statements suggest that some women felt less clear about the impact of this:

> *‘I didn’t really make a birth plan, my labour was led completely [b]y the midwife at the time. I decided it was best to go with the flow and not really make a plan. It was slow and long which ended in an emergency caesarean*.*’*

### COVID related changes to delivery

One of the most notable COVID-related changes reported was the suspension of home births and birthing pools:

> *‘Planned home water birth. All home births cancelled. All water births cancelled’*
>
> *‘Big changes. I was induced, had an epidural […*.*], where I had wanted a water birth. My husband wasn’t allowed to attend until my waters had been manually burst, which did influence me to say yes so that he could join me*.*’*
>
> *‘Husband unable to attend induction or stay with baby & I after birth. Rushed hospital discharge, no visitors, PPE used by staff & I had to wear it whilst in labour too*.

This respondent went on to describe the feeling of being rushed and of having no control:

> *Felt impersonal, rushed, somewhat out of control & birth options v limited (no access to birth centre or home birth). V[sic] different to what we had planned!’*

A number of respondents described being alone, and in one case, the cancellation of plans that had been developed to help prevent the reoccurrence of her postnatal depression:

> *‘No one read my birth plan. I was alone for the majority of my labour. My birth partner was only allowed to join me right at the end*.*’*
>
> *‘I was alone throughout the birthing experience. I couldn’t have a water birth, couldn’t have visitors. Most things planned to help reduce the reoccurrence of postnatal depression could not be put into place*.*’*

One respondent reported having trouble accessing the desired pain relief and assistance during her labour:

> *‘It changed I was induced due to potential infection. And was unable to have the desired pain relief and staffing was low, and they didn’t arrive in my very quick labour’*

These changes and restrictions resulted in some parents feeling considerable distress and anxiety:

> *‘I wasn’t allowed the birth I wanted because of covid. It was hugely traumatic […]’*

### Non-COVID related changes to delivery

Whilst changes to delivery can be expected when giving birth (e.g., *‘I was induced due to potential infection’*) women reported feelings of anxiety and distress with regard to these changes due to the lack of support and communication offered by hospitals:

> *‘My birth plan changed as the baby was in a difficult position but as I was the last in my pregnancy group to give birth and 5/7 of them had had a c-section I was very worried I would have to have a c-section. In hospital it felt like it was my only option’*

These feelings were further exacerbated as a result of COVID-related restrictions to birthing support:

> *‘I would have like[d] a natural labour but my body didn’t go into labour. I suffered from PTSD from my [eldest’s] birth where I was induced therefore it was advised I shouldn’t be induced again. I spent a lot of time worrying about a situation where I might have had to be induced without my husband’s support’*.

Some respondents who described changes to their delivery appeared to adapt well to the changes as a result of feeling supported and informed throughout their journey:

> *‘Things did not go to plan, but I was kept informed, I was consulted on actions taken and my birth plan was considered throughout*.*’*
>
> *‘Change of plan but staff in operation was amazing’*.

In addition to asking families to share their birthing experiences, we asked parents to reflect on the communication they had received prior to the birth of their baby regarding access to birthing partners during the delivery. We identified three key themes related to responses to this question (see *Table 3*, Q7, n = 250): (1) Communication, (2) Impact of fluctuating COVID-related guidance, (3) Anxiety and stress related to changing guidance.

### Communication

The results revealed mixed responses in relation to communication from their hospital prior to the birth of their child. Some parents reported poor communication which added to feelings of anxiety:

> *‘Communication was unclear, causing anxiety. As we have no family in this country, I asked a friend to be a back-up birth partner in case my partner should be barred for such a reason*.*’*
>
> *‘Literally no communication from the hospital so had very little idea what to expect’*

Other parents reported having good communication from their hospital and midwives, in particular noting the use of social media platforms:

> *‘Communication about procedure for spontaneous labour was very clear-used the very helpful midwife - patient liaison Facebook group’*
>
> *‘My local hospital held a webinar with their midwives discussing what to expect at the birth with the new restrictions so I new[sic] exactly what to expect at […]. They also answered any other worries or concerns I had on Facebook messenger. It was really helpful and reassuring*.*’*

### Impact of fluctuating COVID-related guidance

The constant changes to the guidance and restrictions in relation to giving birth during COVID-19 was a major theme within responses related to communication received from the hospital prior to the birth of their child:

> *‘It changed a lot in the build up to birth – as did restrictions on water birth etc. Was very aware that progress could be revoke[d] at any point’*
>
> *‘Things were changing so quickly at the time midwives weren’t 100% sure’*

Many parents related the constant and fast changing nature of guidance as causing feelings of distress:

> *‘The guidelines were changing almost daily. I felt scared and upset. On top of this I was unsure if anyone would be able to look after my son whilst I gave birth*.*’*
>
> *‘Every time I asked the question I was told it could change right up until the morning of my c section-this made me very anxious’*

In addition to the changing rules and guidance, parents highlighted differences between the NHS trusts around COVID guidance and birth:

> *‘As long as a positive test or symptoms aren’t present. It was a concern that they wouldn’t let my partner be present as restrictions were tighter than in other local hospitals’*

### Anxiety and stress related to changing guidance

Many parents reported feelings of anxiety and distress related to not being confident that they would be able to have a birth partner present for the duration of their labour and birth:

> *‘It was awful having no assurance that my partner could attend labour and post-labour. There’s not much else to say except it was the #1 reason for my anxiety in the last few months of pregnancy*.*’*
>
> ‘*I was terrified my whole pregnancy that my husband wouldn’t be able to be there*…*constant source of anxiety waiting for hospitals to update guidance’*
>
> *‘Partner couldn’t come to induction. I found it a really frightening, lonely experience*.*’*

However, the empathy with which this information was conveyed, appeared to have influenced at least one participant’s response to this:

> *‘At one point my midwife told me that I would have to be alone. This was a shock to me and I had a very emotional response. She was also upset by this. I appreciated that this was out of her control and that there was nothing she could do, I just really appreciated her empathic response, I felt less alone in that moment*.*’*

## Discussion

Our study sought to identify the impact of giving birth amidst the changes in public health guidance that were instigated during the COVID-19 pandemic. The CoCoPIP survey provided new mothers with the opportunity to describe their experiences in their own words within the first 6 months following birth. Analyses compared expressed sentiment (i.e., positive, negative and neutral) across lockdown conditions and coded the themes expressed by parents’ open-ended responses, as well as describing the type of delivery, the presence of birthing partners, and changes in birth plan reported by our sample. To date, our study provides the largest sentiment analysis of birth experiences in the UK (Ayers, 2007).

Our results show that 32.6% of participants reported having a CS, of which 47.7% were elective. This represents a significant increase relative to pre-pandemic levels in which around one-quarter of deliveries were typically CS and one-third elective (NHS, 2019). At the start of the pandemic, there were reports that maternal requests for caesarean sections (MRCS) were under a blanket restriction (Romanis & Nelson, 2020). However this subsequently changed, with reports of expectant women in England and Wales opting to have a CS to ensure the presence of birthing partners at birth (Betteley, 2020). This was in response stories of partners being unable to reach the hospital in time for the ‘active labour’ portion of their baby’s delivery (Betteley, 2020). Whilst the current study did not explicitly ask whether respondents elected to have a CS, our results indicated a higher-than-average rate of elective CS, despite the governmental guidance and restrictions in place, suggesting the presence of a partner might have been a potential motivation for this plan.

The results also show that just under half the total sample reported that they were unsure whether their birthing partner would be able to attend the delivery (40.2%), demonstrating uncertainty around access to birth partners throughout the year of the pandemic. When asked to elaborate on how this communication (or lack of) from the hospital made expectant mothers feel (see Q7), a large proportion of individuals reported heightened levels of anxiety and distress. While NHS England has stated that guidance has been clear throughout the pandemic allowing partners to be present for childbirth, this was not always the case (Summers, 2020). For example, the Guardian reported in September 2020 that “three-quarters of NHS trusts are not allowing birth parents to support mothers” (Topping & Duncan, 2020). Within the NHS, each trust was able to issue their own policy, in particular those regarding access to birthing partners (Summers, 2020), leading to inconsistency and confusion across regions and among different expectant families. Our data highlights the way in which the changes in the rules and guidance surrounding birthing preferences and birth partners, not only nationally but between NHS trusts, created confusion and anxiety amongst families. This lack of clear guidance appears to have exacerbated existing feelings of stress and anxiety in women throughout their pregnancy (Aydin, Glasgow, et al., 2021), not only during childbirth.

The results of the sentiment analysis suggest that the fluctuations in guidance and the evolving crisis in terms of the provision of services to pregnant women, led to higher-than-typical (9.3%) reports of negative birth experiences (Rijnders et al., 2008; Smarandache et al., 2016). In our sample, the uncertainty that characterised the initial phase of lockdown seemed to exacerbate the frequency of negative experiences (Smarandache, Kim, Bohr, & Tamim, 2016).

Results from our thematic analysis support those of a survey conducted by Mumsnet and Birthrights between December 2019 – September 2020, which found that many women reported that their decisions with regard to childbirth (e.g., water birth, delayed clamping) were not respected with many reporting their choice was either not considered or disregarded (Mumsnet & Birthrights, 2020). These changes, in addition to uncertainties with regard to access for the birthing partner throughout the pandemic were described as having led to heightened levels of anxiety and a negative childbirth experience.

The findings also suggest, however, that whilst changes to the birth were experienced by a large proportion of our sample, clear communication and support appeared to mitigate these negative childbirth experiences. These findings are consistent with the wider recognition that women’s feelings and ability to exert choice and control during the birth, are more important in terms of long-term wellbeing, than the objective facts of the birth (Cook & Loomis, 2012). It is also now recognised that post-traumatic stress disorder (PTSD) can occur following childbirth and has been found to be influenced by a number of significant factors, including some that were identified by the current study (i.e. negative aspects in staff–mother contact, feelings of loss of control over the situation, and lack of partner support) (Olde et al., 2006). While we do not currently have data with regard to the incidence of PTSD following childbirth during the pandemic, this study found that when families were provided with support and the ability to control the decision-making about the overall birth (e.g., birth plan was followed or communication facilitated by midwives) families concurrently reported a more positive experience with reduced levels of anxiety and stress. Overwhelmingly, however, women reported negative birthing experiences when discussing (i) restrictions in terms of birthing method (i.e., no access to birthing pool or home births), (ii) no offer of support and communication by medical staff and/or (iii) dismissals of their decision with regard to how they wished to give birth.

### Limitations

As data were collected between July 2020 – March 2021, participants experiences reflect a period of fluctuating COVID-related government and healthcare restrictions, from the most severe national lockdown measures to a combination of severe to mild national/local restrictions. Due to the rapidly evolving nature of the governmental guidance related to the pandemic and regional variations in between national lockdowns it, was not possible to collect equal sample sizes at each timepoint. Furthermore, as a result of the fact that this study was conducted as a voluntary online survey, we cannot confirm independently that all responses were by expectant parents or exclude bias in respondents with either positive or negative experience of giving birth. Whilst we advertised this study nationally and specifically worked with national childbirth trusts (NCTs) with an emphasis on areas of low socio-economic status (SES), the majority of participants were white; therefore, the results cannot be generalised to a more ethnically diverse population. Furthermore, while our sample was fairly representative of the UK’s population, there was an underrepresentation of Asian and South Asian parents. This study is part of an ongoing longitudinal study observing the impact of COVID-19 on pregnancy, infant development and parental mental health and we hope we increase the diversity of our sample as recruitment continues. Another limitation is that, of the two questions posed, not every participant gave a response to each one. Finally, from a qualitative perspective, due to the online survey nature of the project, it was not possible to probe by means of interviews, which would for example have helped us further elucidate the links between whether the existence of detailed guidance given by the trust influenced the birth accounts and contributed to the specific reasoning behind the change in birth plan.

### Implications for practice and research

The mitigation measures implemented by the government and the NHS throughout the COVID-19 pandemic have had a significant secondary impact on expectant women and families. Going forward, these findings show the need for clear and consistent guidance to be in place for expectant women giving birth during subsequent lockdowns and any future public health crises. This should include allowances for choice of delivery methods as well as the availability of consistent support for the duration of the labour and birth.

Further research is needed to explore the impact of variation in birth experiences, both on maternal mental health in the postpartum period (Simpson & Catling, 2016), on maternal-infant attachment (Anderson & Cacola, 2017), and on subsequent maternal health and child development (Hernández-Martínez et al., 2020). Maternal recovery and bonding with their infants are particularly salient in light of the relative social isolation experienced by families during the pandemic.

## Conclusions

Changes to birth experiences and offered support-in response to governmental guidance with regard to mitigating the spread of the virus and the increased burden on the healthcare system - has had an adverse effect on the experiences of many pregnant women in England. These findings reinforce the importance of the role of choice and control in women’s childbirth experience. In addition, the findings demonstrate the need to ensure consistent guidance and support to better address the unique health care needs of each pregnant woman in any future lockdowns, as well as the need to observe the potential long-term impact on their offspring.

## Data Availability

Qualitative data generated and analysed during the study will not be made publicly available due to ethical and privacy restrictions, however researchers can submit a research proposal to the Data Sharing Management Committee to request access and collaboration.

## List of abbreviations

NHS: National Health Service
RCM: Royal College of Midwives
CS: Caesarean section
PTSD: Post-traumatic stress disorder

## Declarations

### Ethics approval and consent to participate

Ethics approval for the survey was given by the University of Cambridge, Psychology Research Ethics Committee (PREC) (PRE.2020.077).

## Consent for publication

Not applicable.

## Competing interests

The authors have no potential conflicts of interest to disclose.

## Funding source

This research was funded by a Medical Research Council Programme Grant MR/T003057/1 to MJ, and a UKRI Future Leaders fellowship (grant MR/S018425/1) to SLF. The views expressed are those of the authors and not necessarily those of the MRC or the UKRI. T.A. is supported by the NIHR Cambridge Biomedical Research Centre (BRC). The NIHR Cambridge Biomedical Research Centre (BRC) is a partnership between Cambridge University Hospitals NHS Foundation Trust and the University of Cambridge, funded by the National Institute for Health Research (NIHR), TA is also supported by the NIHR Brain Injury MedTech Co-operative. The views expressed are those of the author(s) and not necessarily those of the NIHR or the Department of Health and Social Care.

## Acknowledgements

We are extremely grateful to all those families who gave their time to participate and to Esther Adememo and Maddie Walton who worked on the CoCoPIP study during their undergraduate studies at Cambridge.

## Author statement

**Ezra Aydin:** Conceptualization, Methodology, Investigation, Data Curation, Formal Analysis, Validation, Writing - Original Draft. **Staci M. Weiss:** Conceptualization, Methodology, Formal Analysis, Visualizing, Writing - Review & Editing. **Zahra Khan:** Investigation, Data Curation, Validation. **Kevin A. Glasgow:** Investigation, Data Curation, Formal Analysis, Writing – Review & Editing. **Topun Austin:** Methodology, Supervision, Writing - Review & Editing. **Mark Johnson:** Supervision, Funding acquisition, Writing - Review & Editing. **Jane Barlow:** Supervision, Writing - Review & Editing. **Sarah Lloyd-Fox:** Conceptualization, Methodology, Supervision, Funding acquisition, Writing - Review & Editing

## Notes

### Competing Interest Statement

The authors have declared no competing interest.

